# The Association of Opioid Use Disorder and COVID-19 in Shahroud, Iran

**DOI:** 10.1101/2021.02.19.21252093

**Authors:** Zhaleh Jamali, Mohammad Hassan Emamian, Hassan Hashemi, Akbar Fotouh

## Abstract

**Background:** COVID-19 quickly spread to the world, causing a pandemic. While some studies have found no link between Opioid Use Disorder (OUD) and COVID-19, the role of the opioid on COVID-19 is challenging. The present study aimed to determine the relationship between OUD and COVID-19.

**Methods:** This was a prospective cohort study. We used data from the third phase of the Shahroud eye cohort study on 4394 participants which started in September 2019 and ended before the COVID-19 epidemic in Shahroud in February 2020. The participants were followed for 10.5 months till November 2020. COVID-19 was detected by RT-PCR on swap samples from the oropharynx and nasopharynx. The incidence of COVID-19 compared in OUD and Non-OUD participants, and relative risk was calculated in Log Binomial Regression model.

**Results:** Among the 4394 participants with a mean age of 61.1 years, 120 people had OUD. The incidence of COVID-19 in participants with OUD and Non-OUD were 3.3% and 4.5%, respectively. The relative risk of OUD for COVID-19 was 0.75 (95% Confidence intervals: 0.28 – 1.98; P= 0.555).

**Conclusions:** Opioid use disorder was not associated with COVID-19. The claim that people with OUD are less likely to develop COVID-19 is not supported by this data.

## Introduction

The COVID-19 pandemic, which began in late 2019 in Wuhan, China, spread rapidly around the world (1). According to the latest report World Health Organization until February 11, 2021, about 107 million people have been infected with COVID-19 worldwide, and more than 2347,000 people have died. During the same period, 1489000 people were infected with COVID-19 and about 59,000 died in Iran (2).

Opioid use disorder (OUD) is a chronic and relapsing disease (3). Opiates are legally prescribed to manage severe and chronic pain, however, some other opiates, such as heroin, opium, and illicit drugs, are abused (4). According to a report by the United Nations Office on Drugs and Crime (UNODC), there are 4 million people with OUDs globally (5). People with OUDs are usually a marginal population that has less access to health care (6). They also suffer from chronic infections, poor physical health, and psychiatric illnesses (7). In these individuals, the risks of hypoxemia from COVID - 19 viral pneumonia, and OUD drug interactions with COVID-19 medications, are significant (8,9). People with OUD need more medical care and because of their greater vulnerability they need more attention in COVID-19 pandemic (10-12). Important risk factors for COVID-19 include old age and underlying diseases such as chronic obstructive lung disease, cardiovascular disease, obesity and diabetes (13). There are a few studies on the relationship between OUD and COVID-19. A study in Iran investigated this relationship in an ecological study and propose a protective role of OUD against COVID-19 (14). This study did not meet the required technical and scientific standards, and their authors retracted it. Insufficient current information regarding relationship between OUD and COVID-19 and cross-sectional studies in this field, urge us to study this relationship in a population-based cohort study.

## Method

Shahroud Eye Cohort Study is a population-based cohort study that has been designed and was launched in 2009 to determine the prevalence and incidence of important eye diseases in adults (40- to 64-year-old) and to identify the risk factors that cause them (15). The second and third phases of this study were conducted in 2014 and 2019. In all three phases, demographic data and past medical and medication histories have been questioned. (15). The data of the present study are from the third phase of this study, which was completed in February 2020 and before the start of the COVID-19 epidemic in Iran. Participants reported any opioid use, including opium and its derivatives, as well as methadone and other narcotics in the interviews. Individuals who consumed any of these substances in any amount were considered OUDs.

A comprehensive system for registration and follow-up of COVID-19 patients has been set up in Shahroud, northeast Iran. All COVID-19 patients, including outpatients and inpatients, were registered in this system (16). Individuals who participated in the cohort study were followed up in the COVID-19 system by using the national identification number for 9 months till the end of November 2020. In this study, a person with COVID-19 has defined as a person who had a positive RT-PCR test in the Nasopharyngeal and Oropharyngeal Swabs samples. The incidence rate of COVID-19 was calculated in two groups of participants (with and without OUD) and then the relative risk of COVID-19 was calculated. Respiratory infection or any suspicion of SARS-CoV-2 infection that led to RT-PCR test for COVID-19 (total positive and negative PCR tests) was investigated as the second outcome. The third outcome was patients who had a positive RT-PCR test or a positive chest CT scan for COVID-19. Descriptive variables were reported by percent and 95% confidence limits and the incidence rates were calculated with Log Binomial Regression.

Everyone in the study participated willingly with written informed consent. Both studies have been approved by the ethics committee of Shahroud University of Medical Sciences.

## Results

In this study, 4394 participants in the third phase of the Shahroud Eye Cohort Study were followed up for 9 months. The mean and standard deviation of the age of the participants was 6.1 ±61.1 years and 59.3% of them were female. Among them, 120 (including 112 men) had OUDs (Table 1).

**Table 1:** The incidences of respiratory infection and COVID-19 by the Opioid user disorders and its relative risks in Log Binomial regression, Shahroud, Iran, 2020

| Opioid User Disorders | Having Respiratory Infection |  |  | RT-PCR positive |  |  | RT-PCR positive or Chest CT scan positive |  |  |
| --- | --- | --- | --- | --- | --- | --- | --- | --- | --- |
|  | No (%) | RR (95% CI) | P value | No (%) | RR (95% CI) | P value | No (%) | RR (95% CI) | P value |
| Yes | 11(9.17) | 1.1 | 0.685 | 4(3.33) | 0.74 | 0.555 | 6(5.00) | 1.04 | 0.908 |
| No | 348 (8.14) | (0.63-1.99) |  | 191(4.47) | (0.28-1.97) |  | 204 (4.77) | (0.47 -2.31) |  |

RR, Relative Risk; CI, Confidence Intervals

The results of Log Binomial Regression showed that the incidence of COVID-19 was 4.47% in the group without OUD and 3.33% in the group with OUD. The relative risk for people with OUD was estimated to be 0.74 (95% CI: 0.28-1.97) which means that the incidence of COVID-19 was not different in the two groups without OUD and with OUD. The results of Log Binomial Regression for two other outcomes including respiratory infection (RI) as well as COVID-19 patients who had positive tests for RT-PCR or chest CT-scan are also shown in Table 1, indicating that OUDs did not have any association with COVID-19.

## Discussion

Opioid use disorder is a common health problem in all societies, and people with OUD are susceptible to infectious diseases in biological, nutritional, behavioral, and quality of life aspects (17). Since the COVID-19 pandemic began, little information has been provided in Iran (18, 19) and sometimes misconceptions have been raised about the resistance of these people to COVID-19. This study showed that the incidence of COVID-19 was not significantly different between people with and without OUD.

In this regard, a study in Iran (20) emphasizes that addicts are more vulnerable to COVID-19 and the odds of mortality in opioid addiction was 3.6 times to other patients. A large study by Wang et al (21) in the United States also highlighted the increased odds of patients with substance use disorders, especially OUDs (adjusted odds ratio = 10.2), to COVID-19. Patients with SUD or OUD were also at higher risk of hospitalization and death (21).

The predisposing factors that can make people with OUD more vulnerable to COVID-19 include nonconformity of social distance, use of shared facilities, lack of personal hygiene, living in crowded groups, and poor physical and mental health (19, 20, 22, 23). Behavioral and economic factors can also increase the risk of COVID-19 among people with SUD (21, 23). COVID-19 can be fatal for people with OUD due to respiratory depression (24) and immune suppression due to long-term substance use (24, 25). This is while, higher doses of opioids use are associated with suppression of the immune system and an increased risk of pneumonia (26). Morphine also increases susceptibility to bacterial and viral infections in injecting drug users (27) which is considered as another mechanism for these people to be susceptible to COVID-19.

On the other hand, some factors may protect OUDs against COVID-19. For example, smoking and tea consumption are more common among people with OUD (28), while there are reports that indicate a resistance to COVID-19 in tobacco (29) and tea (30) consumers. As we know, COVID-19 is transmitted through direct contact of the patient with other people in crowded and closed places (31). Also, drug addicts suffer from mental health and low quality of life and lack of jobs and mental disorders such as antisocial personality, depression, and anxiety (32-34). Therefore, these people are less present in the community and have a less social activity that can reduce the chance of developing COVID-19.

Anyway, it seems that all the predisposing and protective factors neutralize each other’s effects, and the risk of infection with SARS-CoV-2 in people with OUD is similar to other people. Differences in the study age group, type of drug addiction, the presence of other comorbidities and differences in the prevalence of COVID-19 are some of the reasons that the results of this study may differ from other studies.

This prospective study has strengths that include a high sample size, significant follow-up time (9 months), and a well-designed study to record demographic and COVID-19 data. However, this study also had limitations. The absence of distinction between the method of opioid use and the type of addiction is one limitation. Also, the data of this study are limited to the age group of 50 to 74 years, while most opioid users are in the age group of 23 to 44 years (35, 36), which is another limitation of the current study. Although the overall sample size in this study is high, the number of participants with OUD was only 120, and this is another important limitation. The results of the power analysis showed that to identify a 3% difference, with this sample size in the two groups, the study power will be 40%, which is not a significant amount. Thus, further studies with higher sample sizes are suggested in the OUD group.

In summary, people with OUD have a similar risk to the general population for developing COVID-19. Any protection role of OUD against SARS-CoV-2 infection should be discouraged.

## Data Availability

Data will be available on request

